# Pangenome-based identification of cryptic pathogenic variants in undiagnosed rare disease patients

**DOI:** 10.1101/2025.07.08.25330875

**Authors:** Se Song Jang, Seoyeon Kim, Seungbok Lee, Soo Yeon Kim, Jangsup Moon, Jun Kim, Jong-Hee Chae

**Author notes:** To whom correspondence should be addressed., Correspondence may be addressed to Jong-Hee Chae., Correspondence may also be addressed to Jun Kim. S.S.J, S.K, and S.L contributed equally to this work.

## Abstract

**Background:** Despite widespread implementation of exome and genome sequencing, a substantial proportion of rare disease patients remain undiagnosed due to inherent limitations in detecting structural, repetitive, and regulatory variants.

**Methods:** We applied long-read sequencing (LRS) to 40 individuals from 33 previously undiagnosed Korean families. *De novo* assemblies were integrated into a graph-based pangenome workflow, enabling sensitive detection of single-nucleotide, structural, and tandem-repeat variants and direct profiling of CpG methylation.

**Results:** Pathogenic or likely pathogenic variants were identified in 9 (27.3%) families that had remained unsolved despite prior short-read sequencing. The discoveries comprised deep intronic splice-altering SNVs, non-coding regulatory deletions, complex rearrangements, large deletions, tandem repeat expansions, and aberrant methylation profiles. We also implicate *CXXC1* as a novel disease-associated gene, potentially contributing to a global DNA methylation defects, and revealed novel pathogenic variants in established disease genes such as *HEXB* and *NGLY1*, providing insights into underrecognized genetic contributors to rare diseases.

**Conclusions:** LRS coupled with pangenome-based, graph-driven analysis closed a sizable diagnostic gap, broadened the mutational spectra of several Mendelian genes and brought epigenomic evidence into rare disease investigation. These findings support the adoption of long-read, graph-based workflows as a front-line strategy for comprehensive genomic and epigenomic diagnosis.

## Background

Rare diseases affect an estimated 300 million people worldwide and impose physical, emotional, and economic burdens on patients, families, and society (1). Although genomic medicine has advanced rapidly, many individuals whose clinical features strongly suggest a genetic etiology still lack a molecular diagnosis (2, 3). This persistent diagnostic gap hinders applicability of personalized clinical care, accurate prognosis, and enrollment in targeted therapies or clinical trials. Continuous innovation in sequencing technologies and bioinformatic methods is therefore essential to improve diagnostic rates in rare genetic disease.

Conventional short-read sequencing (SRS), including targeted-panel sequencing, whole-exome sequencing (WES), and whole-genome sequencing (WGS), has significantly improved the diagnostic rate of rare genetic disorders (4–9). WES alone achieves a diagnostic yield of roughly 36–41% of cases (10–12). In unsolved patients, adding SR WGS confers a further 7– 19% yield by detecting variant classes that exome approaches often miss, such as structural and deep intronic variants (13, 14). Although short-read (SR) WGS captures a broader range of genomic regions than WES, its SR length limits its ability to resolve structurally complex or repetitive genomic regions.

These challenges have led to increased interest in long-read sequencing (LRS) for rare disease diagnostics (15–28). LRS significantly enhances the detection and characterization of complex structural variants–including large insertions, deletions, inversions, substitutions, and tandem repeat (TR) expansions–as well as variants located within previously unmappable “dark” genomic regions (18, 20, 25, 29–31).

Beyond variant detection, LRS allows haplotype phasing without parental genotyping and supports epigenetic profiling, including DNA methylation or hydroxymethylation (32–34). Moreover, LRS technologies allow the generation of highly contiguous and accurate *de novo* genome assemblies, allowing direct comparison of large haplotype-resolved segments and thus detection of genetic variants of any type and size (35).

The recent emergence of population-specific genome assemblies through global pangenome initiatives is providing valuable background variation data, improving filtering and prioritization of candidate variants. Projects such as the Human Pangenome Reference Consortium (HPRC), Korean Pangenome Project, Chinese Pangenome Project, Arab Pangenome Project, Emirates Pangenome Project and others are contributing to a more representative understanding of human genomic diversity (36–40). However, as rare disease diagnosis requires extensive control datasets, there is still a need for high-quality and population-specific datasets.

In this study, we applied LRS and pangenome analysis to resolve undiagnosed Korean patients with rare diseases. These patients had previously undergone SR WES and WGS with available parental or other familial data but remained nonetheless undiagnosed. Genome assembly and variant discovery were performed on 40 patients from 33 unrelated families (Fig. 1). We further provide genome-wide methylation profiles and Korean-specific LR (long-read) genome datasets as publicly accessible resources. These results highlight the diagnostic value of assembly-based LRS and its potential to elucidate the complex genetic architecture underlying rare diseases.

**Fig. 1:**
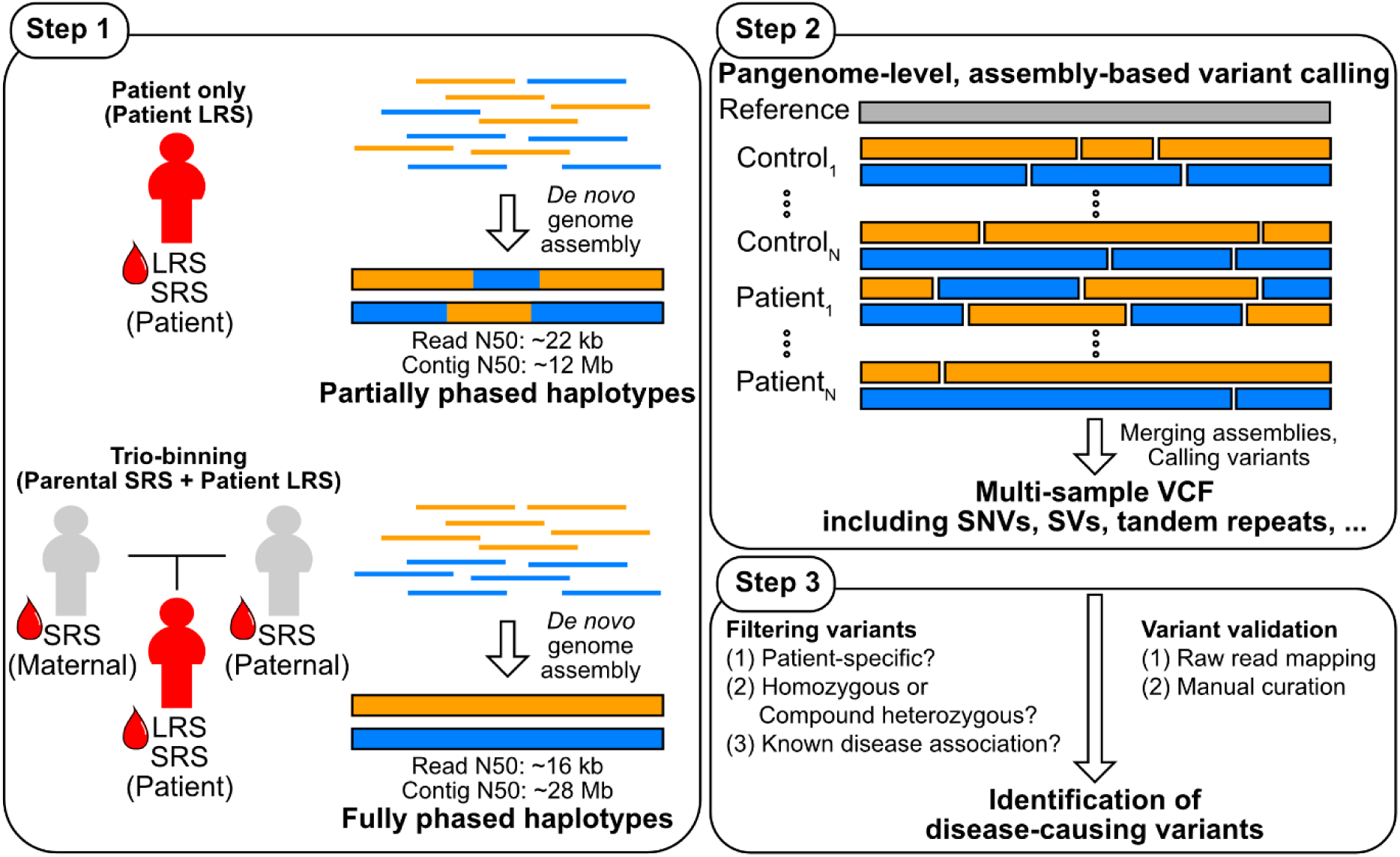
Schematic overview of pangenome-based variant discovery and interpretation workflow. *Step 1.* Overview of sequencing strategies and genome assembly approaches. For patient-only long-read sequencing (LRS), partially phased assemblies were generated. For selected trios or quartets, trio-binning was applied using parental short-read sequencing (SRS) and patient LRS data to obtain fully phased haplotypes. *Step 2.* Pangenome-level, assembly-based variant calling using multiple reference genomes (GRCh38 and T2T-CHM13) and graph-based alignment enabled comprehensive detection of structural variants (SVs), single nucleotide variants (SNVs), and tandem repeat expansions. HPRC genome assemblies were used as control cases. Epigenetic features were investigated using raw base-calling data. *Step 3.* Patient-specific variants were filtered based on their allelic and known disease association information. Filtered variants were manually assessed using raw read validation and known gene and variant databases. This workflow facilitated identification and interpretation of disease-associated variants in previously undiagnosed rare disease patients.

## Methods

### Patient recruitment and sample collection

The patients enrolled in this study exhibited clinical features suggestive of rare genetic disorders and remained undiagnosed despite extensive prior genetic testing, including WES, WGS, and, in some cases, RNA sequencing of muscle tissue obtained via biopsy (Fig. S1).

Participants were recruited through the Department of Genomic Medicine at Seoul National University Hospital, and phenotypic data were reviewed by experienced clinicians. LRS was pursued in cases with persistent diagnostic uncertainty and available biospecimens from the patient and, where possible, parents or affected siblings. Clinical characteristics and sample metadata are summarized in Table S1.

### Whole-genome sequencing and *de novo* genome assembly

SR WGS data of trios was produced using the Illumina NovaSeq6000 platform. LR WGS data from patients and available siblings were generated using one of the following LRS platforms: Pacific Biosciences (PacBio) Sequel IIe, PacBio Revio, or Oxford Nanopore Technologies (ONT) PromethION 24. Detailed sequencing metadata of each sample is shown in Table S1. Read lengths and quality scores of LRS datasets were summarized using bioawk (version 20110810; *bioawk-c fastx’{print length($seq), meanqual($seq)}’*) and SeqKit stats (version 2.8.0; *seqkit stat-a*) (https://github.com/lh3/bioawk) (41, 42).

For samples without parental sequencing data, PacBio long reads were assembled into contig using Hifiasm (version 0.19.8-r603; default parameters) (43–45). For samples with available parental SR WGS data, parental *k*-mers were first calculated for the SR WGS data using yak (version 0.1; *yak count-b 37*) (https://github.com/lh3/yak). Based on these parental *k*-mers and the patients’ HiFi reads, a pair of fully phased genome assemblies was produced for each sample using Hifiasm (version 0.19.8-r603; *hifiasm-1 paternal.yak-2 maternal.yak HiFi.fastq*) (43–45). For samples sequenced using the ONT platform, raw reads were polished using Herro (version model_R10_v0.1; default parameters); polished reads were assembled using Hifiasm (version 0.19.8-r603; default parameters) (43–46).

Then, the assembly quality value (QV) was calculated by first generating a *k*-mer database from each sample’s SR WGS data with Meryl (version 1.3; *meryl k=21 count*) before evaluating it with Merqury (version 1.3; *merqury.sh*) (47). For samples with available parental SR WGS data, Hap-mer was additionally used to construct paternal-and maternal-specific *k*-mer databases, enabling the assessment of haplotype-level completeness (version 1.3; *hapmers.sh*) (47). Contig length metrics for each *de novo* assembly were obtained using SeqKit (version 2.8.0; *seqkit stats*) and bioawk (version 20110810; *bioawk-c fastx’{print length($seq)}’*) (https://github.com/lh3/bioawk).

### Graph-based pangenome construction for variant calling and discovery of candidate causal variants

A pangenome graph was built using the Minigraph-Cactus pipeline by integrating GRCh38.p14, T2T-CHM13 v2.0, HPRC Year 1 assemblies, and our newly generated genome assemblies (version 2.9.2; *cactus-pangenome--reference GRCh38 CHM13--giraffe clip filter--vcf--viz--odgi--chrom-vg clip filter--chrom-og--gbz clip filter full--gfa clip full--vcf--giraffe--gfa--gbz--chrom-vg*) (36, 48, 49). During graph construction, 22 assemblies from the low-depth PacBio 10× group (11 individuals) did not integrate cleanly enough to support a high-confidence graph and were therefore excluded from the build. The pipeline’s final output—a multiallelic VCF—served as input for variant discovery in patients who remained genetically unsolved after SR WGS analysis.

Assuming that conventional small variants were addressed during the preceding SR WGS analysis, this step concentrated on loci whose longest allele was ≥ 3 bp. Allele lengths at each locus were standardized using Scikit-learn’s StandardScaler and grouped by hierarchical clustering using AgglomerativeClustering (version 1.5.2; *AgglomerativeClustering(distance_threshold=1, n_clusters=None)*), so that alleles of identical or similar length fell into the same cluster. Cluster groups containing any allele from the HPRC sample were filtered out. Among the remaining patient-specific clusters, we focused on two patterns: (i) clusters composed exclusively of siblings and (ii) clusters in which a single patient harbored a length-distinct allele absent from all other individuals. Then, each candidate allele was inspected in raw read alignments to confirm the length difference and the gene disrupted by the variant evaluated with respect to the patient’s clinical presentation. Variants whose predicted gene disruption plausibly accounted for the observed phenotype were retained as final, curated causal candidates.

To compute the pangenome growth curve, we produced a base-pair-level cumulative growth curve without GRCh38 reference paths using Panacus (version 0.3.3; *panacus ordered-histgrowth-c bp-l 1,2,1,1,1-q 0,0,1,0.5,0.1-S-O* and *panacus-visualize*) (50).

### Identification of 5-methylcytosine sites and gene-level DNA methylation analysis

PacBio HiFi reads were aligned to the GRCh38 reference genome using pbmm2 (version1.13.1; *pbmm2 align--preset HIFI--sort*) (50, 51). Methylation ratios at CpG sites were subsequently obtained using pb-CpG-tools (version 2.3.2; *aligned_bam_to_cpg_scores--model pileup_calling_model.v1.tflite*) (https://github.com/PacificBiosciences/pb-CpG-tools).

For ONT reads, alignment and sorting to GRCh38 were performed with Dorado (version 0.6.2; *dorado basecaller hac* with a hac model dna_r10.4.1_e8.2_400bps_hac@v4.2.0) and SAMtools (version 1.20; *samtools sort*), respectively (Oxford Nanopore Technologies, 2023; https://github.com/nanoporetech/dorado) (52). Methylation detection and analysis were performed using the ONT wf-human-variation pipeline (version 2.1.0; *wf-human-variation--snv--sv--str--mod--cnv--sex--tr_bed--use_qdnaseq--qdnaseq_bin_size 50--basecaller_cfg dna_r10.4.1_e8.2_400bps_hac@v4.2.0*) (https://github.com/epi2me-labs/wf-human-variation). Methylation ratios were extracted from the resulting bedmethyl files using the following filtering criteria: coverage between 20–200× and modification score > 75%. The identified CpG sites as well as their methylation ratios were classified into defined genomic regions (intergenic region, upstream, promoter, 5’ UTR, CDS, intron, and 3’ UTR). The genomic coordinates of these regions were based on Ensembl’s evidence-based annotation of GRCh38.p14 (version 46; Ensembl 112), except for the upstream and promoter regions, which were defined separately due to lacking annotations: The upstream region was defined as 1–5 kb upstream of the gene start site and the promoter region as that within 1 kb immediately upstream of the gene start site. Finally, the number of CpG sites and the average methylation ratio for each gene region were extracted for each sample.

### Identification of tandem repeats and detection of pathogenic repeat expansions

TR loci were delineated across the GRCh38 reference genome using Tandem Repeat Finder (version 4.09; *trf 2 5 7 80 10 50 2000-l 6-h*) (53). Each TR was assigned to its precise genic context—intergenic, upstream, promoter, 5′ UTR, CDS, intron, or 3′ UTR— to produce the full list of repeat alleles for every gene present in GRCh38.

Population-level TR variants were extracted from the intersection between these TR coordinates and the multiallelic VCF generated with the Minigraph-Cactus pangenome pipeline using BCFtools (version 1.17; *bcftools view*) (52). The resulting TR variant calls were projected onto reference alleles with BCFtools (version 1.17; *bcftools consensus*) to yield the complete set of repeat alleles for each TR locus present in the cohort population (52).

To isolate candidate pathogenic expansions, alleles were filtered in four sequential steps. First, only alleles unique to our patient cohort were retained. Second, within each locus group, the allele with the maximum repeat count was selected. Third, an expansion index was calculated for every allele as 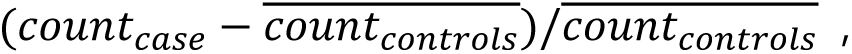 where an index of 1 denotes a two-fold expansion relative to controls. Fourth, alleles exhibiting an expansion index ≥ 1 for both the maximum repeat count and the total repeat length (bp) were considered candidate pathogenic repeat expansions (e.g., for the allele (CAG)_11_(CAT)(CAG)_14_, the maximum repeat count equals 14 due to (CAG)_14_ and the total repeat length equals 78 bp by calculating the length sum of (CAG)_11_, (CAT), and (CAG)_14_, namely 3×11+3+3×14). We further checked if these filtered candidate TR loci were contained within OMIM-listed gene regions. Finally, these final candidate TR calls near disease-associated genes were subjected to manual curation: we evaluated concordance with each patient’s clinical phenotype and confirmed the repeat-length expansion by visual inspection of raw read alignments.

## Results

### Long-read data enable high-quality *de novo* genome assembly sufficient for accurate variant detection

High-quality LRS data were successfully obtained for all enrolled individuals. We used PacBio and ONT LRS platforms to sequence 28 and 12 individuals, respectively. We obtained an average coverage of 11.95× and 31.29× for PacBio samples (hereafter, PacBio 10× and 30× samples, respectively) and 35.67× for ONT samples and mean read length of 14.85 kb, 15.53 kb, and 14.21 kb for PacBio 10×, PacBio 30×, and ONT samples (Table S2). For patients with or without parental SR WGS data, long reads were assembled into contigs using LRS-only or trio-binning approaches, respectively. Although LRS-only assemblies were only partially phased while trio-binned assemblies achieved full phasing, all assemblies exhibited sufficient contiguity (N50: PacBio 10× samples, 0.25–3.50 Mb; PacBio 30× samples, 10.78–59.97 Mb; ONT samples, 1.43–58.08 Mb) to enable phasing and comparison of large genomic segments (Fig. 2A and Table S3).

**Fig. 2:**
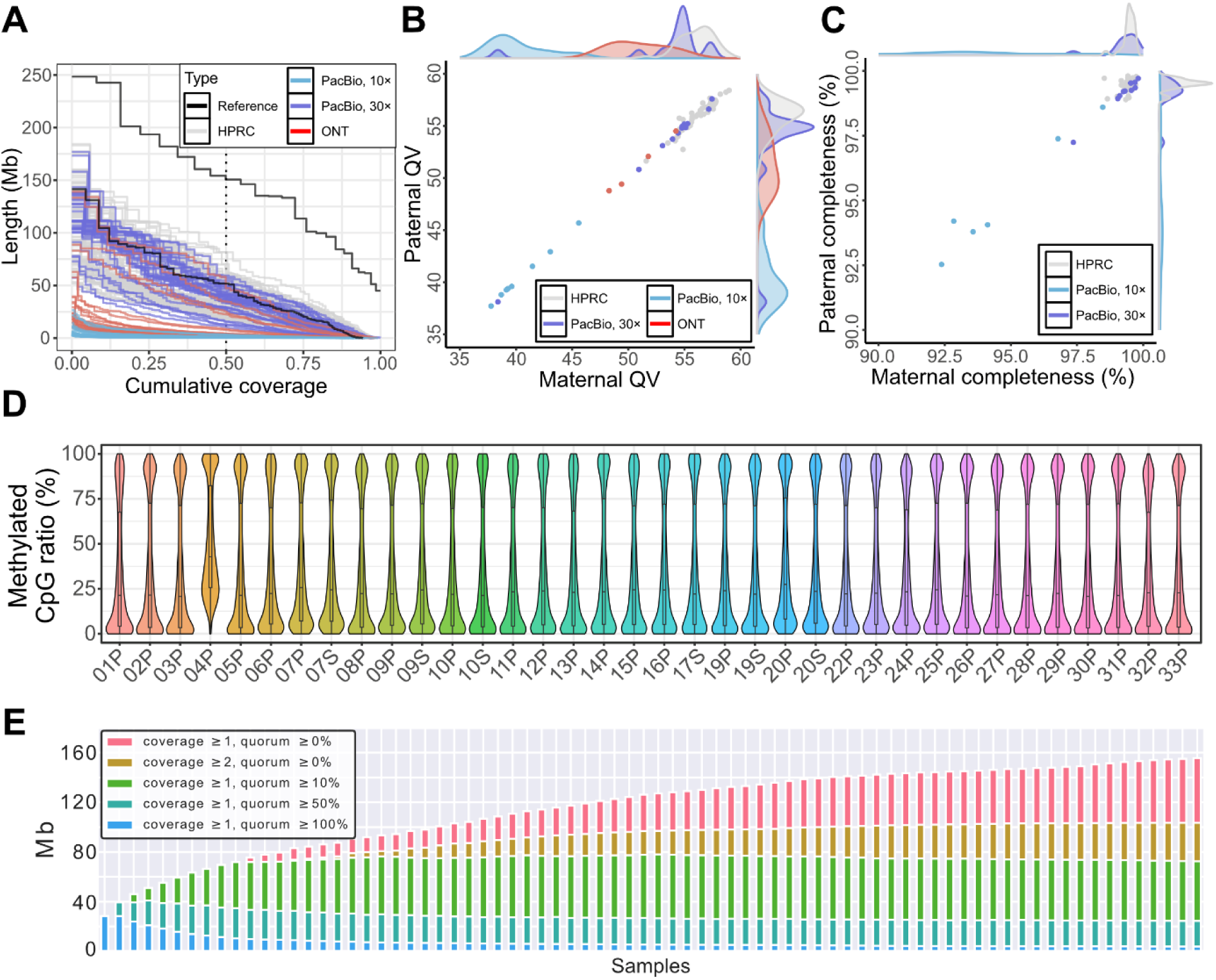
Quality assessment of LRS-generated genome assemblies. **A** Cumulative contig length distributions for each assembly, stratified by sequencing platform and coverage depth. Assemblies generated at ≥ 30× coverage exhibit contiguity comparable to reference-grade genomes (HPRC samples). **B** Comparison of maternal and paternal assembly quality values (QV) calculated by *k*-mer distributions of assembly and raw SRS data. **C** Phasing completeness of maternal and paternal haplotypes, demonstrating near-complete assemblies for trio-binned samples and strong performance across high-coverage datasets. **D** Average methylated CpG ratios in promoter regions for cohort samples. Our sample prefix SNUH-is not shown. As presented in Fig. S2, SNUH-17P, SNUH-18P, SNUH-18S, and SNUH-21P samples originated from fibroblasts instead of blood. **E** Pangenome growth curves using HPRC and our cohort samples. Coverage indicates the number of haplotypes sharing variants and quorum the number of haplotypes sharing variants. For example, coverage ≥ 1 and quorum ≥ 100% indicates that variants are shared by all sample assemblies, while coverage ≥ 1 and quorum ≥ 0% represent singleton variants.

All genome assemblies were high-quality in terms of base-and phasing-level accuracy and our LRS data exhibited consistent methylation profiles (Fig. 2). For samples with available LRS and SRS, we calculated base-level accuracy of the genome assembly by comparing *k*-mer distributions of matched genome assembly and SR WGS data. All genome assemblies exhibited 37.72–57.59 quality values (QVs), specifically, QV37.40–45.69 for PacBio 10×, QV38.12–57.59 for PacBio 30×, and QV48.29–54.51 for ONT samples (Table S4). Of these, genome assemblies constructed using PacBio 30× and ONT samples were comparable to state-of-the-art HPRC genome assemblies (QV51.59–58.98 for HPRC samples; Fig. 2B).

For samples where both maternal and paternal SR WGS were available, we also assessed the phasing-level of the genome assembly by analyzing haplotype-specific *k*-mer distributions between each haplotype genome assembly and each parental SR WGS data. These samples were solely sequenced using PacBio HiFi platforms; they exhibited 92.38–99.81% haplotype-specific *k*-mer completeness, indicating that the haplotype genome assemblies of each sample were accurately phased (Fig. 2C). Specifically, PacBio 10× and 30× samples exhibited 92.38–98.60% and 97.24–99.81% *k*-mer completeness, while HPRC did 98.53–99.80% *k*-mer completeness (Table S4). Further, the methylation profiles of these samples were highly consistent among samples, except for the patient with triple X syndrome (SNUH-04P), which showed outlier methylation patterns (Fig. 2D for blood samples and Fig. S2 for fibroblast samples).

In these patients, we analyzed variants by integrating genome assemblies into a single pangenome graph. For this pangenome graph, we merged 94 and 58 genome assemblies of 47 HPRC samples and our 29 cohort samples, respectively, excluding 22 genome assemblies of 11 PacBio 10× samples due to low depth (Fig. 2E). This pangenome graph contains a total of 20.5M SNVs, 73-K SVs, and 573-K TR loci. Moreover, our genome assemblies revealed patient-specific variants, consisting of 1.9M SNVs, 29 K SVs, and 159 TR expansion loci. Next, these patient-specific variants were investigated to identify potential causal variants in undiagnosed patient samples (Fig. 1 and Table 1). LRS enabled detection of complex structural variants, noncoding deletions, repeat expansions associated with methylation changes, and accurate phasing of compound heterozygous variants, many unresolved by previous SRS methods.

**Table 1:** Summary of identified variants and diagnostic contributions of long-read sequencing in undiagnosed rare disease patients.

| Sample ID | Genetic Diagnosis | Gene | Variant Information | Zygosity | LRS Contribution |
| --- | --- | --- | --- | --- | --- |
| **SNUH-01P** | Congenital myopathy-7A (OMIM: 608358) | *MYH7* | Missense variant-exon 31 (NM_000257.4:c.4325T>C;p. Leu1442Pro) | Het | Initially detected by SRS and confirmed by LRS |
| **SNUH-02P** | ReNU syndrome (OMIM: 620851) | *RNU4-2* | 1-bp insertion-exon 1 (NR_003137.3:n.64_65insT) | Het | Initially detected by SRS and confirmed by LRS |
| **SNUH-03P** | MRD74 (OMIM: 620688) | *HNRNPC* | 27-bp deletion-exon 8 (NM_001025108.2:c.850_876del;p. Arg284_Asp292del) | Het | Initially detected by SRS and confirmed by LRS |
| **SNUH-04P** | Triple X syndrome with *CXXC1*-related epigenetic disorder | *CXXC1* | Nonsense variant-exon 8 (NM_014593.4:c.1011dupT;p. Glu338Ter) | Het | Identified a methylation outlier pattern |
|  |  |  | 47 XXX, methylation outlier (global methylation defects) |  |  |
| **SNUH-05P** | KINSSHIP syndrome (OMIM: 619297) | *AFF3* | (CGG)<sub>145</sub> repeat expansion-promoter (XM_005263943.5:c.-199_-222GCG[145]) | Het | Identified 145-repeat expansion and hypermethylation in the promoter region |
|  |  |  | Hypermethylation in the promoter region of the *AFF3* gene |  |  |
| **SNUH-06P** | Congenital disorder of deglycosylation-1 (OMIM: 615273) | *NGLY1* | Missense variant-exon 6 (NM_018297.4:c.925T>C;p. Cys309Arg) | Het | Identified a novel SV and confirmed *trans* configuration |
|  |  |  | 16,588-bp deletion-exon 1, 2 (NM_018297.4:c.-5705_246+6066del) | Het |  |
| **SNUH-07\*** | Sandhoff disease (OMIM: 268800) | *HEXB* | Deep intronic variant-intron 6 (NM_000521.4:c.771+985G>A) | Het | Identified a novel SV and confirmed *trans* configuration |
|  |  |  | 6,875-bp deletion-exon 4, 5 (NM_000521.4:c.512-1201_670-2853del) | Het |  |
| **SNUH-08P** | Cockayne syndrome type A (OMIM: 216400) | *ERCC8* | 18,756-bp deletion and 7-bp insertion-exon 10 (NM_000082.4:c.400-3403_1041+1368delinsAATAGAA;p. Thr134_Gln347del) | Het | Identified both SVs and confirmed *trans* configuration |
|  |  |  | Exon 4 rearrangement in *ERCC8* | Het |  |
| **SNUH-09\*** | NEDCASB (OMIM: 619121) | *SHMT2* | Missense variant-exon 8 (NM_005412.6:c.977G>T;p. Gly326Val) | Het | Phasing confirmed without parental sample |
|  |  |  | Missense variant-exon 9 (NM_005412.6:c.1075C>T;p. Arg359Trp) | Het |  |
| **SNUH-10\*** | Allan-Herndon-Dudley syndrome (OMIM: 300523) | *SLC16A2* | 2,876-bp deletion-intron 1 (NM_006517.5:c.430+38618_430+41493del) | Hemi | Detected a noncoding deletion undetected by SR WES/WGS |
| **SNUH-11P** | Neuronal intranuclear inclusion disease (OMIM: 603472) | *NOTCH2NLC* | (GGC)<sub>94</sub> repeat expansion-promoter (chr1:g.149390802_149390829GGC[94]) | Het | Identified 94-repeat expansion |
| **SNUH-12P** | MDDGA4 (OMIM: 253800) | *FKTN* | 3,072-bp insertion-exon 10 (3' UTR) (NM_006731.2:c.*4392_*4393ins) | Hom | Identified the precise breakpoint |

\*Patient and affected sibling shared the same variant.
MRD74, autosomal dominant intellectual developmental disorder-74; NEDCASB, neurodevelopmental disorder with cardiomyopathy, spasticity, and brain
abnormalities; MDDGA4, Muscular dystrophy-dystroglycanopathy with brain and eye anomalies (type A4); UTR, untranslated region; Het, heterozygous; Hom,
homozygous; LRS, long-read sequencing; SRS, short-read sequencing; SV, structural variant WES, whole-exome sequencing; WGS, whole-genome sequencing.

### Our pangenome analysis outperforms previous short-read-based approaches

To systematically assess the diagnostic contribution of LRS, we analyzed a cohort of 33 previously undiagnosed rare disease families. Sample IDs (SNUH-01 to SNUH-33) were assigned in order of diagnostic outcome: newly diagnosed by both SRS and LRS (SNUH-01 to SNUH-03), newly diagnosed by LRS (SNUH-04 to SNUH-12), and undiagnosed despite LRS (SNUH-13 to SNUH-33). Table 1 summarizes clinical features, causative genes, variant types, and phasing information for all diagnosed families.

A molecular diagnosis was established in 12 of 33 previously undiagnosed families. Of these, 3 cases (SNUH-01 to 03) harbored pathogenic variants retrospectively detectable by SR WGS and included to assess the confirmatory capacity of LRS. The remaining 9 diagnoses (SNUH-04 to 12) were achieved exclusively through LRS-based variant detection or phasing. Among the nine LRS-contributed cases, the identified diagnostic variants included deep intronic SNVs, large deletions, repeat expansions with associated methylation abnormalities, and complex rearrangements. In several cases, LRS enabled phasing of compound heterozygous variants even in the absence of parental samples. These findings underscore the broad utility of LRS for detecting complex and noncoding variants in unresolved rare disease cases.

### A triple X syndrome patient with a *de novo CXXC1* SNV exhibits a global methylation defect signature

A female patient with a prior diagnosis of triple X syndrome (SNUH-04P) presented with profound global developmental delay, microcephaly (<3rd percentile), and syndromic facial features. She was unable to walk independently, and her speech was limited to a few simple words such as’mom’ and’dad,’ with no ability to form sentences. This unusually severe phenotype could not be sufficiently explained by triple X syndrome alone. LRS-based genome-wide methylation profiling revealed a markedly altered epigenetic signature, including higher promoter methylation and an abnormal 5′ UTR methylation pattern across all chromosomes (Fig. 2D and Figs. S2 and S3). Retrospective re-analysis of trio-based exome sequencing identified a *de novo* one-base-pair frameshift insertion in *CXXC1* (c.1011dupT; p. Glu338Ter), a gene involved in H3K4 methylation and maintenance of DNA methylation homeostasis (54–57). This variant was absent from both the gnomAD (v4.1.0) and Bravo databases (https://bravo.sph.umich.edu/). In addition, *CXXC1* has a probability of loss-of-function intolerance (pLI) score of 1.0, indicating a high likelihood of haploinsufficiency.

Together, these findings support the hypothesis that *CXXC1* dysfunction plays a causal role in the patient’s atypical phenotype, highlighting the diagnostic potential of methylation profiling in rare diseases where sequence-level variants are absent or uninformative.

### A tandem repeat expansion in *AFF3* is associated with promoter hypermethylation

A patient with neurodevelopmental delay (SNUH-05P) harbored a (CGG)_145_ TR expansion in the promoter region of *AFF3*, identified by LRS (Fig. 3A). The expansion was associated with promoter hypermethylation, consistent with previously reported cases of *AFF3*-related intellectual disability (Figs. 3B and 3C) (11, 58). This repeat expansion was undetected in prior SRS analysis due to its size and location within a GC-rich region. This case demonstrates the value of LRS in detecting unstable TRs with epigenetic consequences.

**Fig. 3:**
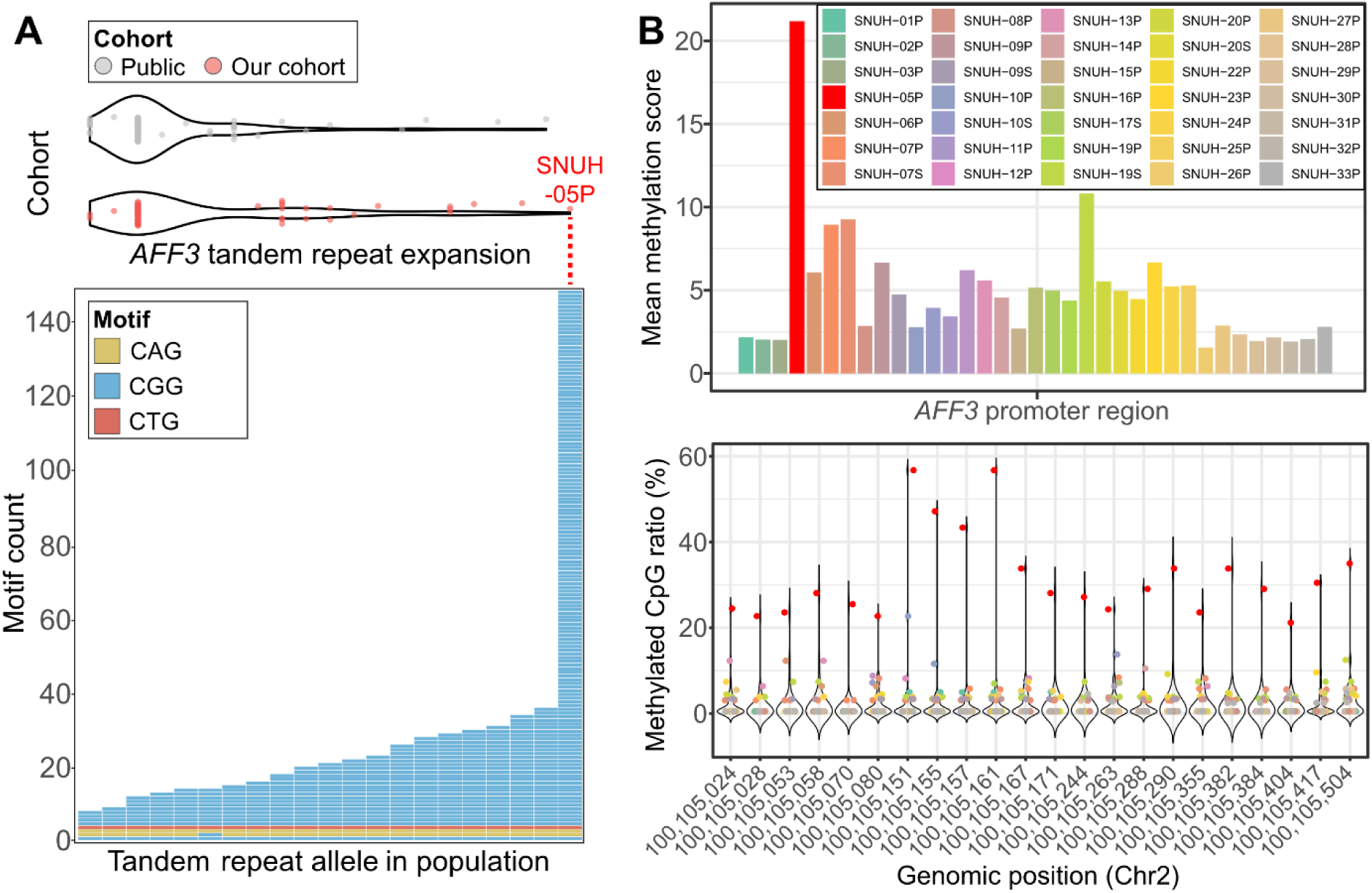
Tandem repeat expansion and hypermethylation profile in *AFF3*. **A** Tandem repeat motif counts and allele frequency distributions in HPRC and our cohort. The exceptional TR expansion in SNUH-05P is denoted as large stacks of CGG motifs. **B** Hypermethylated patterns in the promoter (upper) and specific CpG sites where SNUH-05P exhibited higher methylation ratios (lower).

### Phased genome assemblies reveal compound heterozygous variants in *NGLY1*, *HEXB*, and *ERCC8*

A patient with features consistent with a congenital disorder of deglycosylation (SNUH-06P) remained undiagnosed despite trio-based WES and WGS. LRS identified compound heterozygous variants in *NGLY1*, a gene associated with an autosomal recessive congenital disorder of deglycosylation-1 (Fig. 4A) (59). We identified a missense variant (*NGLY1*: c.925T>C; p. Cys309Arg), classified as a variant of uncertain significance (VUS) in ClinVar (Variation ID: VCV000219542.5) but extremely rare (gnomAD exome allele frequency (AF) = 1.73×10^−7^; genome AF = 6.57×10^−6^), with a high Combined Annotation Dependent Depletion (CADD) score of 26.3 and strong evolutionary conservation (PhyloP100 = 9.021). The second variant was a previously unreported 16.6 kb deletion spanning exons 1–2.

**Fig. 4:**
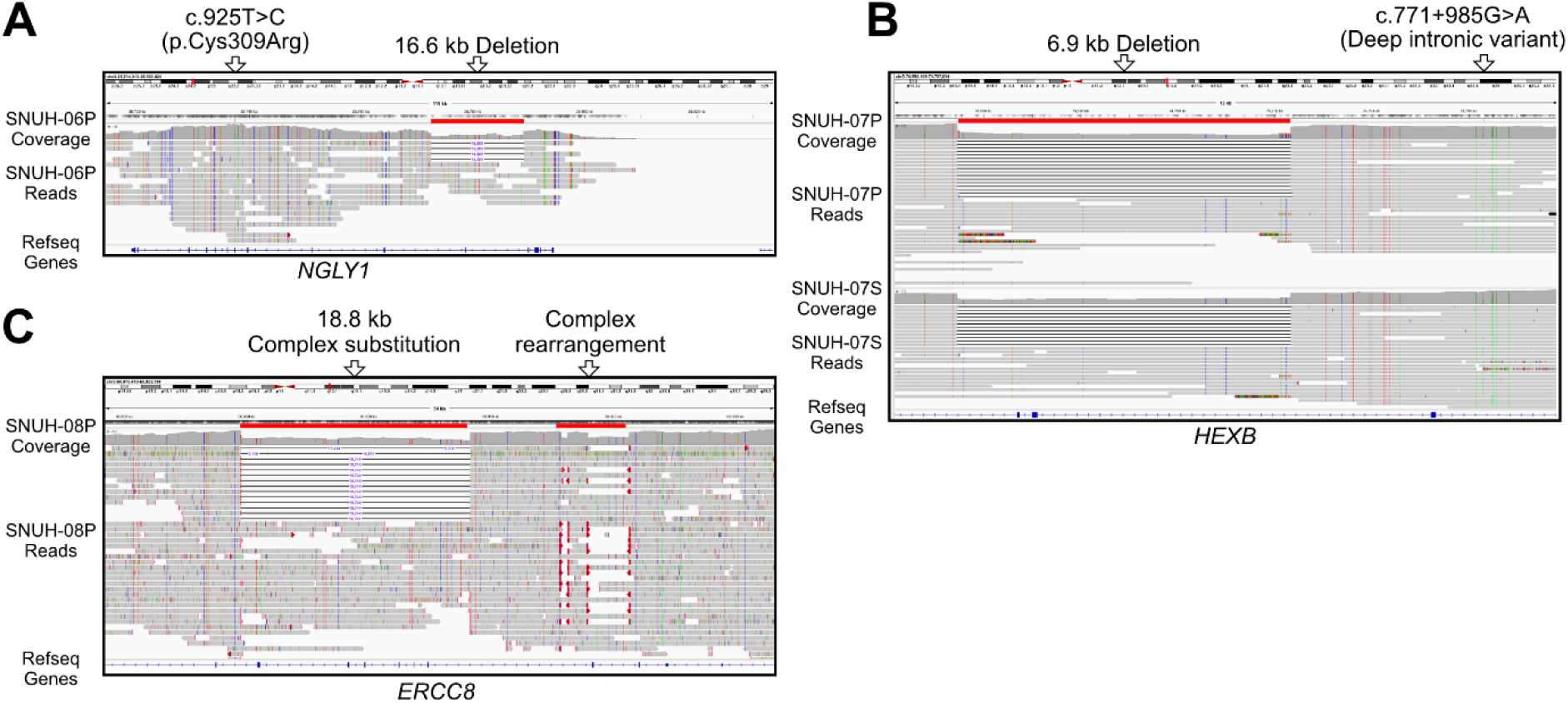
Representative cases demonstrating the diagnostic contributions of LRS. **A** Complex structural variant including a large deletion and insertion in *NGLY1* (SNUH-06P). **B** Compound heterozygous deep intronic SNV and exon deletion in *HEXB* (SNUH-07 Siblings). **C** Compound heterozygous complex rearrangement and complex substitution in *ERCC8* (SNUH-08P).

Two siblings presenting with a progressive motor disorder and features clinically resembling neuronal ceroid lipofuscinosis (SNUH-07P and SNUH-07S) were found to carry compound heterozygous variants in *HEXB*, which causes Sandhoff disease, a condition with partially overlapping clinical manifestations (60, 61). LRS identified an approximately 6.9 kb deletion spanning exons 4–5 *in trans* with a deep intronic SNV (*HEXB*: c.771+985G>A; Fig. 4B). The intronic variant missed by WES due to its noncoding location, is extremely rare (gnomAD AF = 1.31×10^−5^), predicted to alter splicing as an acceptor gain (SpliceAI score = 0.85), and has a high CADD score = 25.2. LRS allowed for phasing and structural resolution, highlighting its utility in detecting and characterizing compound heterozygous variants involving both coding and noncoding regions.

Two siblings (SNUH-08P and SNUH-08S) presented with failure to thrive, short stature, deep-set eyes, nystagmus, episodic encephalopathy, spastic-ataxic gait, and dental crowding, and they were clinically suspected to have Cockayne syndrome. Brain MRI showed hypomyelination and vermian hypoplasia. LRS revealed a complex rearrangement involving inversion and insertion within *ERCC8*, in a previously reported region associated with Cockayne syndrome type A (62). Additionally, we identified a novel complex substitution of 18.8 kb by 7 bp in *ERCC8* (Fig. 4C). LRS also enabled phasing of the two variants, confirming location on opposite alleles (*in trans*) without parental testing. PCR amplification and Sanger sequencing validated the result (Fig. S4). This case illustrates the utility of LRS for detecting and phasing both known and novel structural variants in autosomal recessive Mendelian disorders.

### Resolution of previously ambiguous pathogenic variants through LRS-based phasing, assembly, and repeat quantification

Next, we reexamined previously reported pathogenic variants to resolve phasing and structural ambiguities using LRS. In SNUH-09 siblings, despite the absence of a parental sample, LRS confirmed that the *SHMT2* variants were *in trans* (Supplementary Text Case 1). In the SNUH-10 siblings and SNUH-12P, LRS clarified the structure and breakpoints of unresolved deletions and insertions in *SLC16A2* and *FKTN*, respectively (Supplementary Texts Case 2 and Case 3) (63). In SNUH-11P, we previously identified pathogenic *NOTCH2NLC* GGC repeat expansions, with repeat sizes estimated of 82–90 (64). With our current assembly-based LRS approach, we narrowed down the repeat size to 94, demonstrating the enhanced resolution of this *de novo* assembly. Together, these cases highlight the advantages of LRS in phasing, structural resolution, and repeat quantification—capabilities that enhance diagnostic precision beyond the limits of SRS.

### Potential repair mechanisms for large variants based on genomic signatures

We further annotated the potential repair mechanisms underlying the formation of these newly found large SVs by analyzing DNA repair signatures in their flanking sequences. For the 16,588-bp deletion in *NGLY1*, we found 1-bp microhomologous sequences at the breakpoints. This microhomology implies that a DNA double-strand break (DSB) was repaired by polymerase theta-mediated end-joining (TMEJ), which excises both DSB ends annealling them to repair the DSB site (Fig. 5A). Similarly, the 6,875-bp and 2,876-bp deletions in *HEXB* and *SLC16A2*, respectively, exhibited ∼300-bp long homologous sequences. This longer homology implies that the single-strand annealing (SSA) repair mechanism requiring much longer homologous sequences to anneal both DSB ends than TMEJ acted on the corresponding DSB sites (Figs. 5B and 5C). The complex substitution of 18,756 bp by 7 bp in *ERCC8* exhibited several microhomologous sequences, supporting that TMEJ was involved in this complex substitution, as this type of templated insertion could be generated solely by polymerase theta. In this case, TMEJ may have repaired the DSB sites through several rounds of template switching and microhomology search followed by synthesis (Fig. 5D).

**Fig. 5:**
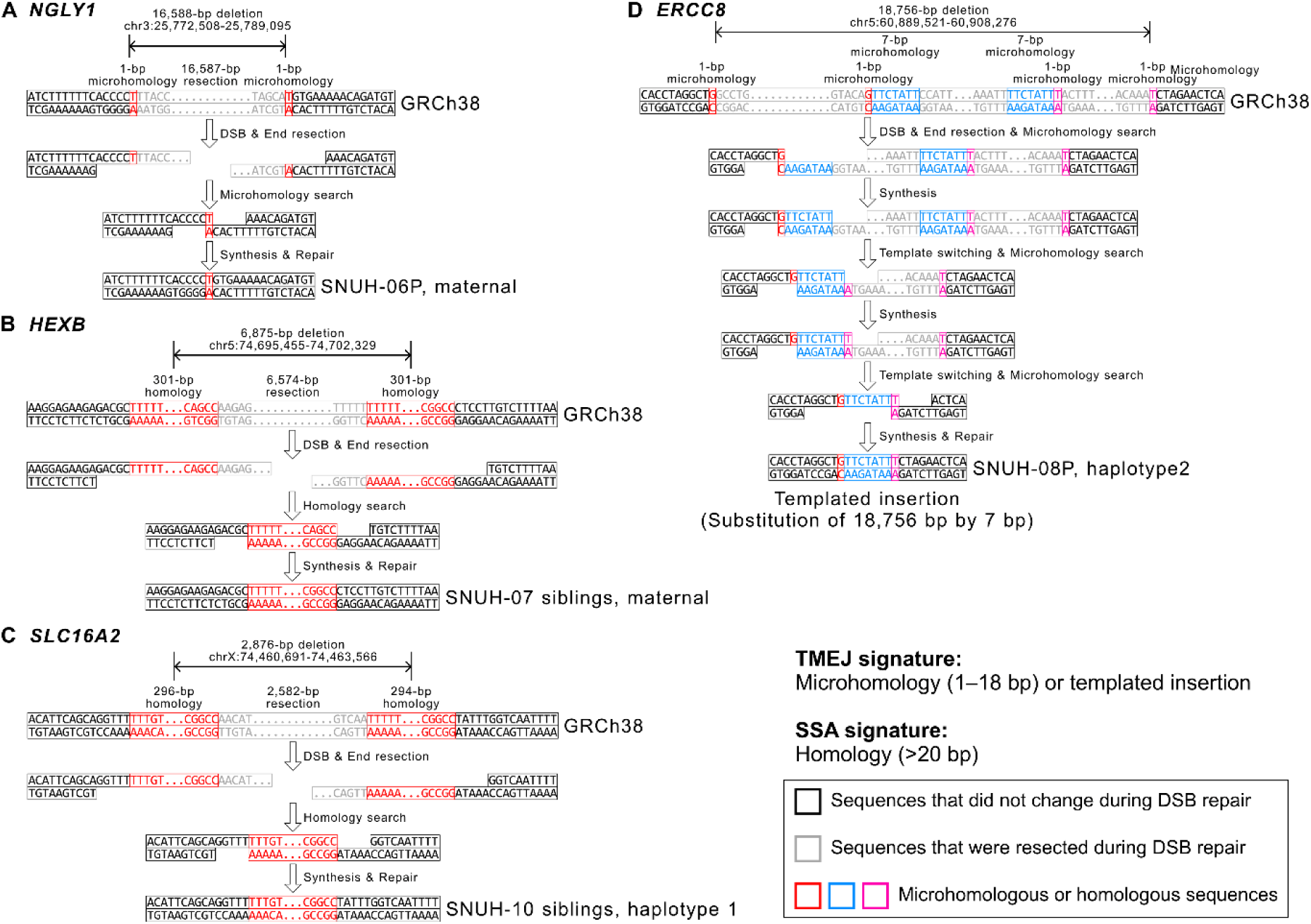
Annotating potential repair mechanisms underlying newly identified large SVs. **A** 16,588-bp deletion in *NGLY1*. A DSB may have occurred in the gene as both DSB ends were resected to expose microhomologous sequences. These microhomologous sequences may have been annealed and repaired (a signature of TMEJ). **B**–**C** 6,875-bp and 2,876-bp deletions in *HEXB* and *SLC16A2*, respectively. Although similar events may have occurred in these genes, their homologous sequences were much longer (∼300-bp), which is a signature of SSA rather than TMEJ. **D** Complex substitution of 18,756 bp by 7 bp in *ERCC8*. Three rounds of template switching and microhomology search followed by synthesis are shown. Double-stranded DNA sequences are indicated; single-stranded DNA sequences represent resected DNA molecules exposing the microhomologous or homologous sequences to be annealed. Same colors represent a pair of microhomologous or homologous sequences. DSB, double-strand break; TMEJ, polymerase theta-mediated end-joining; SSA, single-strand annealing.

## Discussion

LRS combined with assembly-and graph-based genome analysis reconstructs patient-specific haplotypes and directly compares them to a pangenome reference graph, providing a robust framework for uncovering variants often missed by SRS. Through this approach, our study delivered molecular diagnoses of approximately 30% of families who had exhausted conventional testing, demonstrating that sequence length remains a critical determinant of diagnostic success.

By integrating *de novo* assembly with graph-based variant calling, we not only increased variant detection sensitivity but also achieved phasing of compound heterozygous variants without requiring parental data. This capability is particularly valuable for adult patients or isolated cases where trio sequencing is not feasible. In addition, LRS unlocked access to variant classes elusive to conventional SRS, while simultaneously enabling methylation profiling. Representative cases in our cohort illustrate the distinct diagnostic power of LRS: for example, the resolution of a novel complex substitution of 19 kb by 7 bp in *ERCC8*, the discovery of a promoter repeat expansion in *AFF3* with concurrent hypermethylation, and the identification of a *CXXC1* variant associated with a genome-wide methylation outlier profile.

For *AFF3*, several studies have demonstrated that single nucleotide variants, TR expansions, and methylation changes in its promoter region are associated with intellectual disability (11, 58, 65, 66). To date, only two studies have reported both a TR expansion and associated promoter methylation changes at the same *AFF3* locus as in our patient–one using pyrosequencing in three families, and the other employing LRS in two trios (11, 58). Our study represents the third report supporting these observations, and is the first in an East Asian population. Despite the lack of targeted therapies, the epigenetic nature of this mechanism suggests potential avenues for therapeutic development.

This study provides new evidence implicating *CXXC1* in disease through globally aberrant DNA methylation. To date, *CXXC1* has not been directly associated with human disease. However, it forms a histone H3 lysine 4 (H3K4) methyltransferase complex together with *SETD1A* that can regulate both H3K4 and DNA methylation (67–69). *SETD1A* is a well-characterized OMIM gene involved in neurodevelopmental disorders (OMIM #618832 and #619056) (69–73). *CXXC1* can directly bind unmethylated CpG sites and recruit a H3K4 methyltransferase, thereby blocking the access of DNA methyltransferases to these CpG sites and inhibiting DNA methylation (69, 74–80). This function provides a plausible explanation for the global DNA methylation defects observed in the patient harboring said mutation.

Although further validation is necessary, our findings provide strong evidence that *CXXC1* may be a critical mediator of global DNA methylation dysregulation in neurodevelopmental disorders.

Notably, we identified novel large variants in *NGLY1*, *HEXB*, and *ERCC8* and a noncoding regulatory deletion in *SLC16A2*, expanding the known mutational spectrum of these genes. Identifying diverse variants is especially critical to maximize diagnostic yield, given the growing availability of targeted therapies, as such findings may point to clinically actionable or potentially treatable targets. For example, *HEXB* mutations cause Sandhoff disease, for which substrate reduction and gene therapies are currently in development (81–87). In the *SLC16A2*-related Allan-Herndon-Dudley syndrome, early intervention with thyroid hormone analogs has clinical benefits, and recent preclinical studies further support the therapeutic potential of antisense oligonucleotides and AAV-based gene therapy to restore brain thyroid hormone transport (88–90). *NGLY1* deficiency, though ultra-rare, has become a focus of emerging therapeutic strategies, including microbial metabolic therapy, pharmacological chaperones, and gene transfer approaches. Recent studies also suggest metabolic and proteostatic vulnerabilities that may offer additional intervention points (91–94). These findings accentuate how LRS and pangenome-based analysis not only improve diagnostic yield but also play an important role in guiding therapeutic decisions, particularly in the years to come.

Moreover, precise breakpoint resolution afforded by LRS allowed us to infer the DNA damage repair mechanisms underlying these structural changes. Micro-or homologous sequences near these large deletions were genomic signatures of homology-directed repair mechanisms. It specifically implies that SSA was the potential mechanism of forming the deletions in *HEXB* and *SLC16A2* and TMEJ was for the deletion and complex substitution in *NGLY1* and *ERCC8*, respectively. Interrogating the potential repair mechanisms of pathogenic variants provides valuable insight into how these variant alterations have emerged and persisted in the human population.

By generating high-quality genome assemblies, methylation profiles, and variant catalogs from individuals of Korean ancestry, we also contribute population-specific resources that can support future variant filtering and discovery efforts. Although functional validation was beyond our scope, the convergence of genomic and phenotypic evidence supports the clinical relevance of our findings. Nonetheless, >50% patients in our cohort remain undiagnosed, highlighting the need for broader application of LRS and continued advances in analytical methods.

## CONCLUSIONS

Our integrative LRS and pangenome-based analysis demonstrates that direct interrogation of both the genome and epigenome can uncover pathogenic variations that elude detection by conventional SRS. The discovery of *CXXC1* as a novel disease gene, along with expanded allelic spectra in established genes (*NGLY1*, *HEXB*, *ERCC8*, and *SLC16A2*), underscores the diagnostic value of LRS technologies. This study also supports the integration of epigenomic insights into precision medicine workflows. Broader application of this framework may improve diagnostic yield and accelerate the translation of comprehensive genomic and epigenomic data into actionable clinical care.

## Data Availability

The raw sequencing data generated during the current study are available in the NCBI BioProject database, https://www.ncbi.nlm.nih.gov/bioproject/, under the accession number PRJNA1284003.

## LIST OF ABBREVIATIONS

LRS: Long-read sequencing
SRS: Short-read sequencing
WES: Whole-exome sequencing
WGS: Whole-genome sequencing
SR: Short-read
TR: tandem repeat
HPRC: Human Pangenome Reference Consortium
LR: Long-read
SV: Structural variant
SNV: Single nucleotide variant
PacBio: Pacific Biosciences
ONT: Oxford Nanopore Technologies
QV: Quality value
pLI: Probability of loss-of-function intolerance
VUS: Variant of uncertain significance
AF: Allele frequency
DSB: DNA double-strand break
TMEJ: Polymerase theta-mediated end-joining
SSA: Single-strand annealing

## SUPPLEMENTARY INFORMATION

Additional file1. Fig. S1: Overview of long-read sequencing datasets for individual patients. Additional file2. Table. S1: Clinical characteristics and sample metadata of individuals included in this study.

Additional file3. Fig. S2: Global methylation profiles categorized by genic components.

Additional file4. Table. S2: Summary statistics of long-read sequencing data from 40 patients.

Additional file5. Table. S3: Summary statistics of genome assemblies from 40 patients.

Additional file6. Table. S4: Quality value and completeness of haplotype-resolved genome assemblies for each patient.

Additional file7. Fig. S3: Methylation profiles across chromosomes of the patient (SNUH-04P) harboring a *CXXC1* mutation.

Additional file8. Fig. S4: Validation of a novel substitution of SNUH-08P identified in *ERCC8*. Additional file9. Supplementary Text Case1–3: Diagnosed cases using LRS in addition to previous SRS analyses.

## DECLARATIONS

### Ethics approval and consent to participate

All participants (or their legal guardians) provided written informed consent for study participation. The study protocol was approved by the Institutional Review Board (IRB number: 2402-149-1518) of Seoul National University Hospital.

### Consent for publication

Written informed consent for publication was obtained from all patients or their legal guardians. The consent permits the publication of relevant clinical information while explicitly prohibiting the disclosure of personally identifiable information.

### Availability of data and materials

The raw sequencing data generated during the current study are available in the NCBI BioProject database, https://www.ncbi.nlm.nih.gov/bioproject/, under the accession number PRJNA1284003. The 94 pangenome assemblies of HPRC used during the current study are available in the Zenodo repository at https://zenodo.org/record/5826274/files/HPRC-yr1.agc?download=1 (HPRC-yr1.agc for HPRC Year 1 genome assemblies).

### Competing interests

The authors declare that they have no competing interests.

## Funding

SNUH Lee Kun-hee Child Cancer & Rare Disease Project, Republic of Korea [22B-001-0100]. National Research Foundation of Korea (NRF) grant funded by the Korea government (MSIT) [2025-00519278]. Funding for open access charge: Not Determined.

### Author’s contributions

S.S.J.: Conceptualization, Methodology, Formal analysis, Investigation, Writing—original draft, Writing—review & editing. S.K.: Methodology, Formal analysis, Investigation, Writing— original draft, Writing—review & editing. S.L.: Conceptualization, Methodology, Investigation, Writing—original draft, Writing—review & editing, Supervision. S.Y.K.: Investigation, Writing—review & editing. J.M.: Investigation, Writing—review & editing. J.K.: Conceptualization, Methodology, Writing—original draft, Writing—review & editing, Funding acquisition, Supervision. J-H.C.: Conceptualization, Methodology, Funding acquisition, Supervision. All authors read and approved the final manuscript.

## Acknowledgements

We acknowledge and thank all the participating individuals and their families.

## Notes

### Competing Interest Statement

The authors have declared no competing interest.

### Author Declarations

The study protocol was approved by the Institutional Review Board (IRB number: 2402-149-1518) of Seoul National University Hospital.

