## Supplementary Figures for "Pangenome-based identification of cryptic pathogenic variants in undiagnosed rare disease patients"

**SUPPLEMENTARY FIGURE**

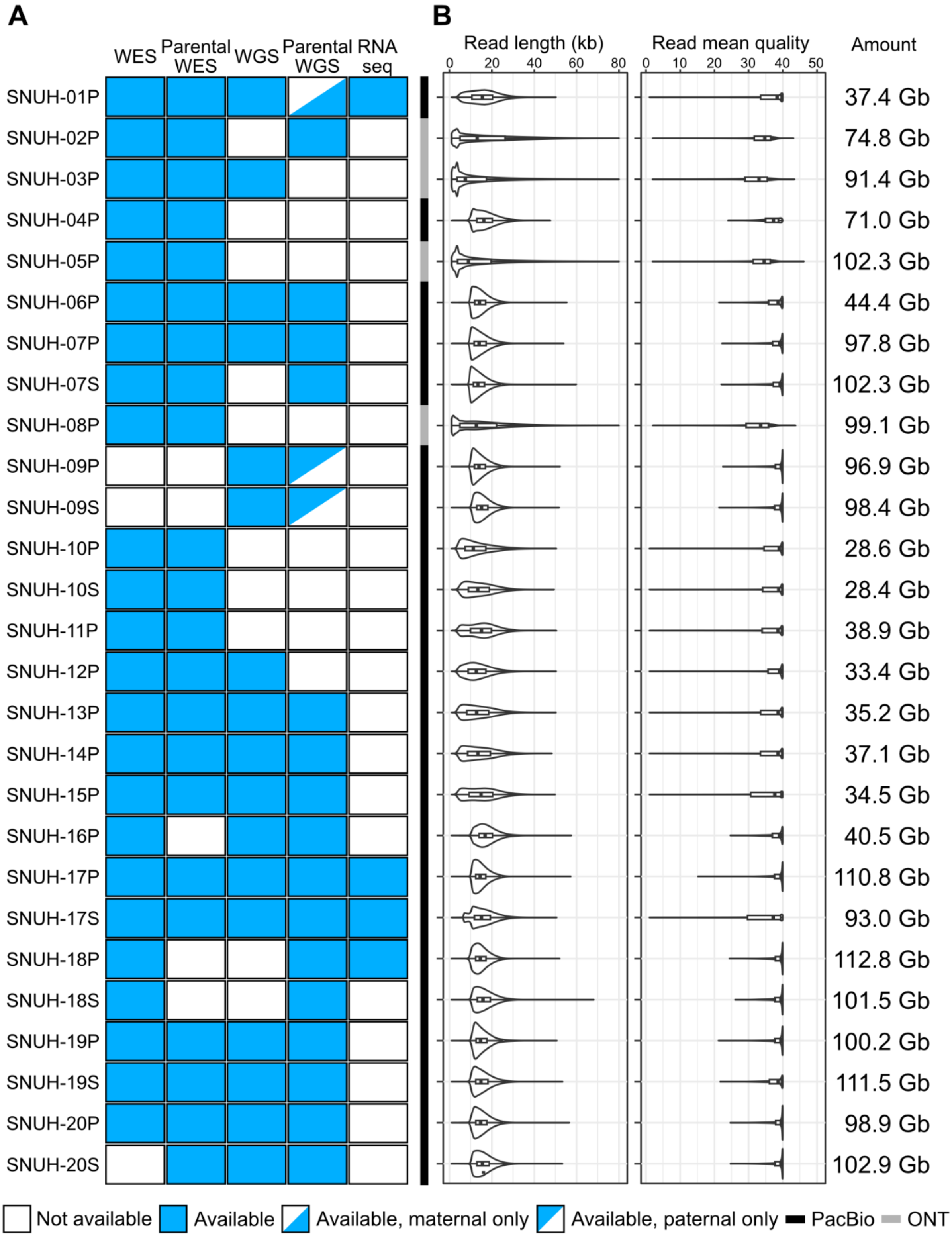

(Continued from previous page)

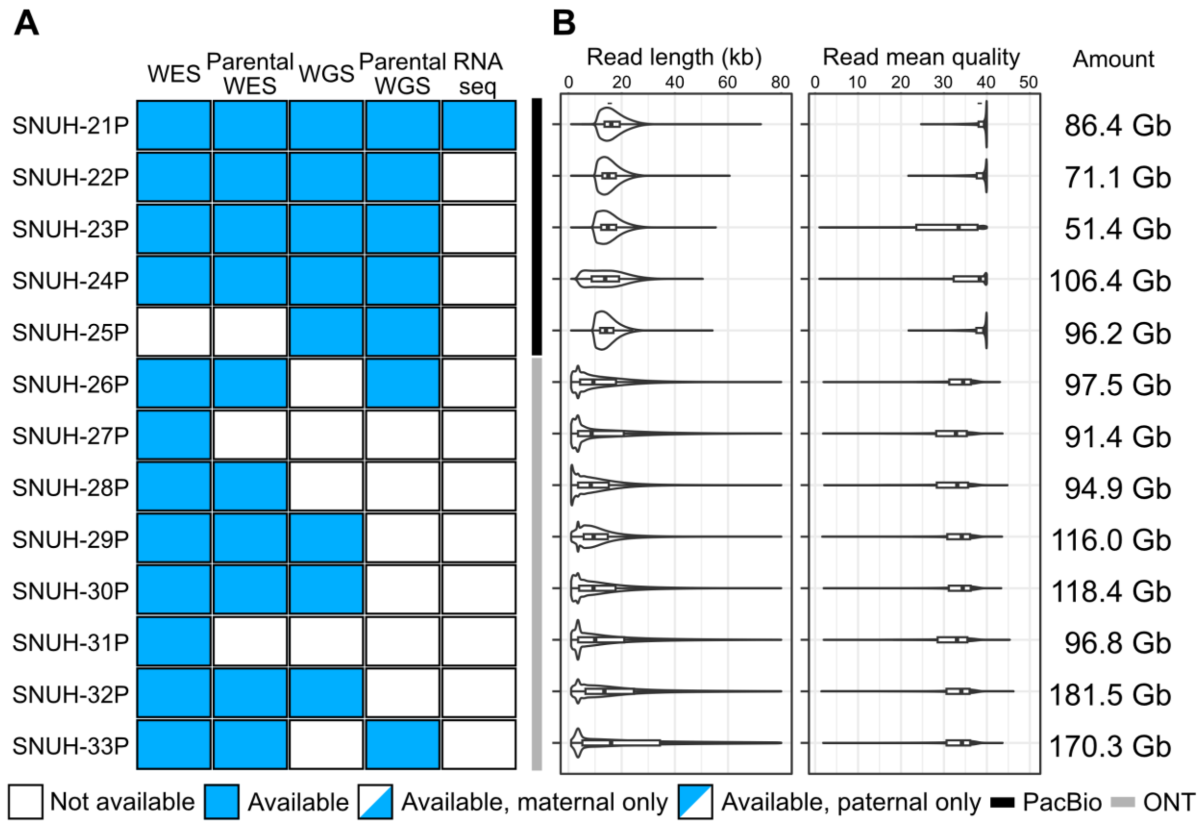


**Figure S1.** Overview of long-read sequencing datasets for individual patients.

**A** Available omics datasets (WES, WGS, RNA-seq) for each sample. Blue boxes indicate availability of existing SRS data. **B** LRS statistics, including read length distribution, average read quality, and total base yield. Violin plots with box plots show the distribution of read lengths and mean read quality.

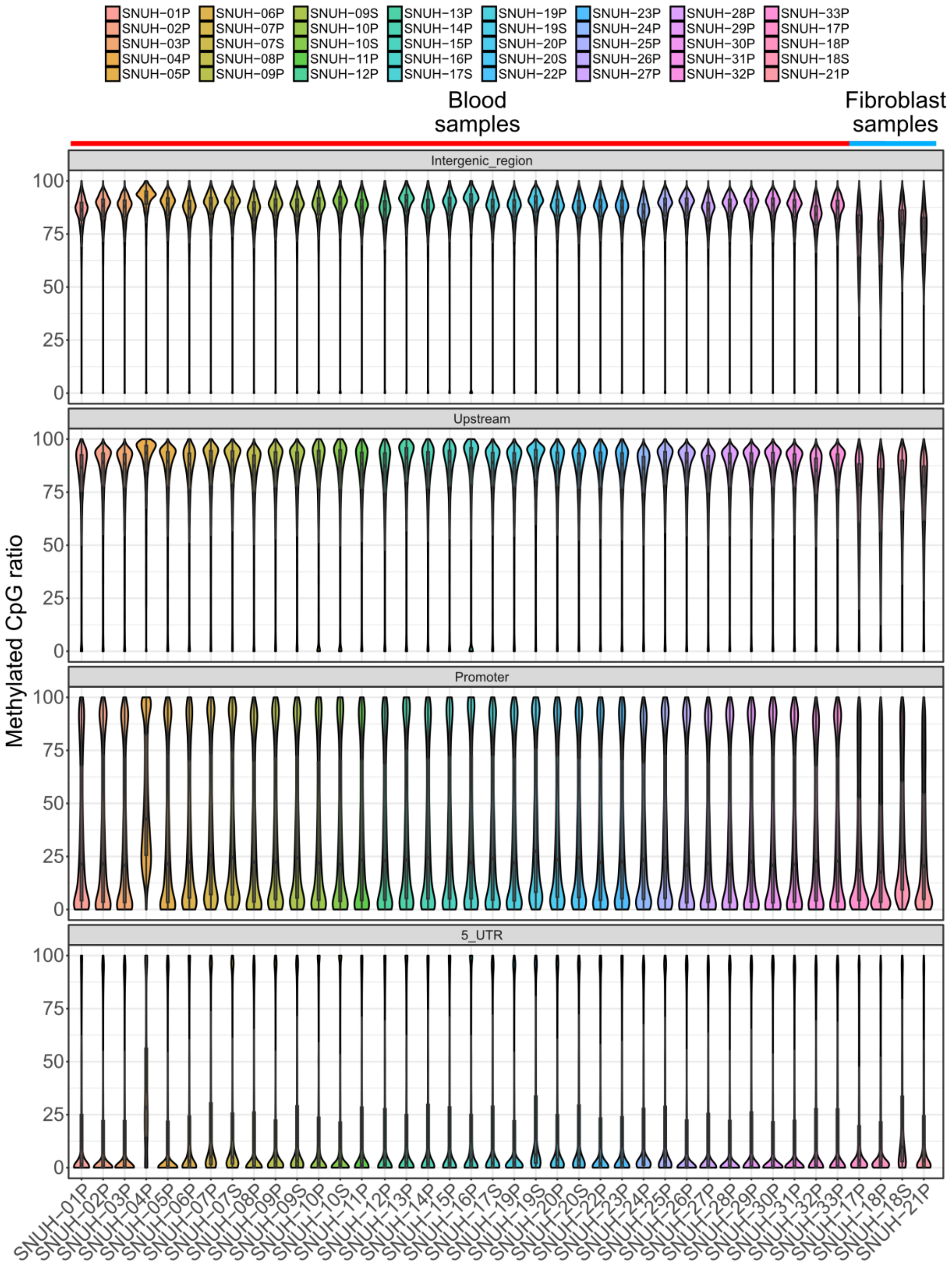
SNUH−01P
SNUH−02P
SNUH−03P
SNUH−04P
SNUH−05P
SNUH−06P
SNUH−07P
SNUH−07S
SNUH−08P
SNUH−09P
SNUH−09S
SNUH−10P
SNUH−10S
SNUH−11P
SNUH−12P
SNUH−13P
SNUH−14P
SNUH−15P
SNUH−16P
SNUH−17S
SNUH−19P
SNUH−19S
SNUH−20P
SNUH−20S
SNUH−22P
SNUH−23P
SNUH−24P
SNUH−25P
SNUH−26P
SNUH−27P
SNUH−28P
SNUH−29P
SNUH−30P
SNUH−31P
SNUH−32P
SNUH−33P
SNUH−17P
SNUH−18P
SNUH−18S
SNUH−21P
Blood samples
Fibroblast samples
Intergenic_region
Upstream
Promoter
5_UTR
Methylated CpG ratio
100
75
50
25
0

(Continued from previous page)

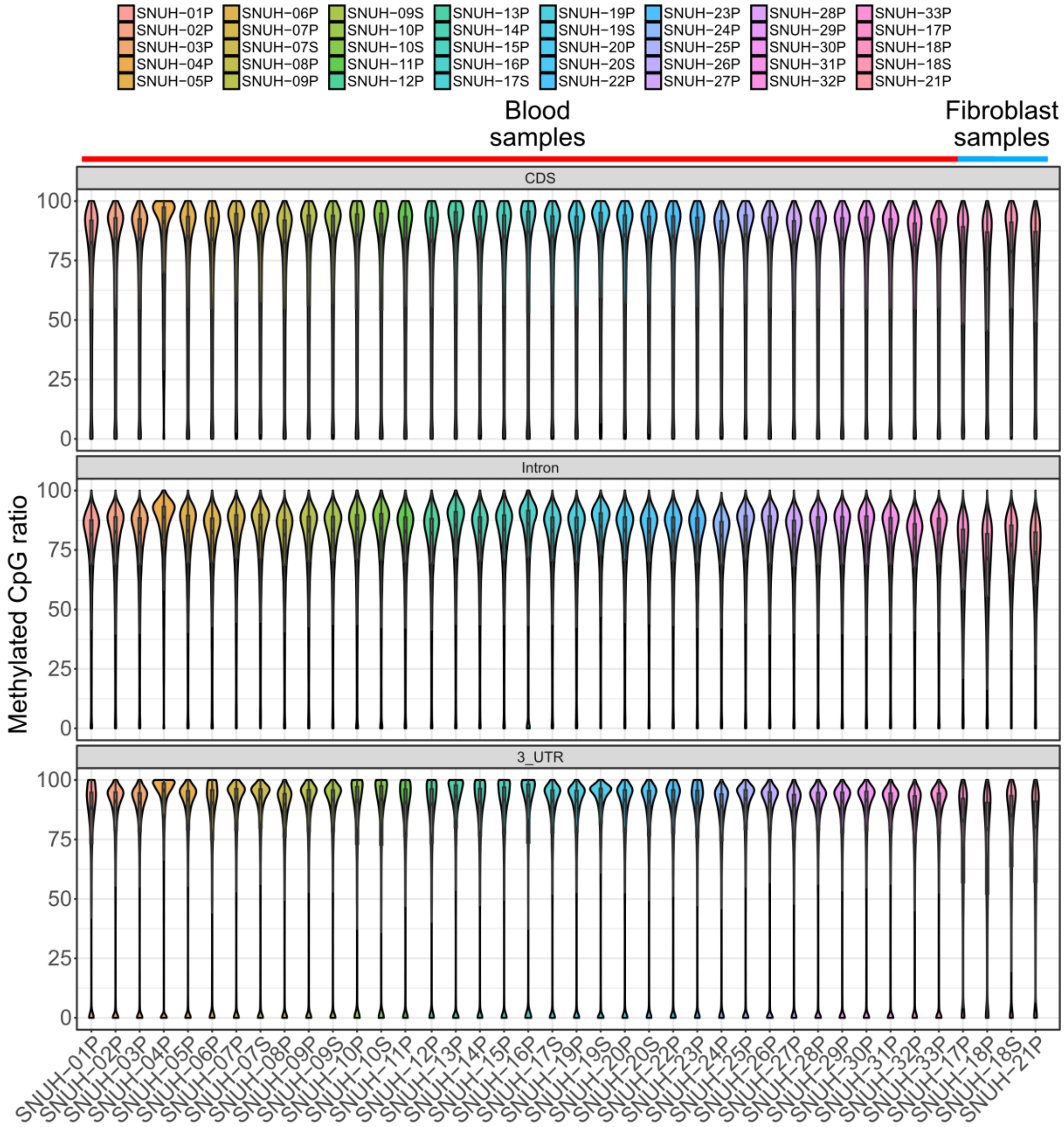


**Figure S2.** Global methylation profiles categorized by genic components.

Methylated cytosine ratios across intergenic regions, upstream regions, promoters, 5′ untranslated regions (UTRs), coding sequences (CDS), introns, and 3′ UTRs are shown.

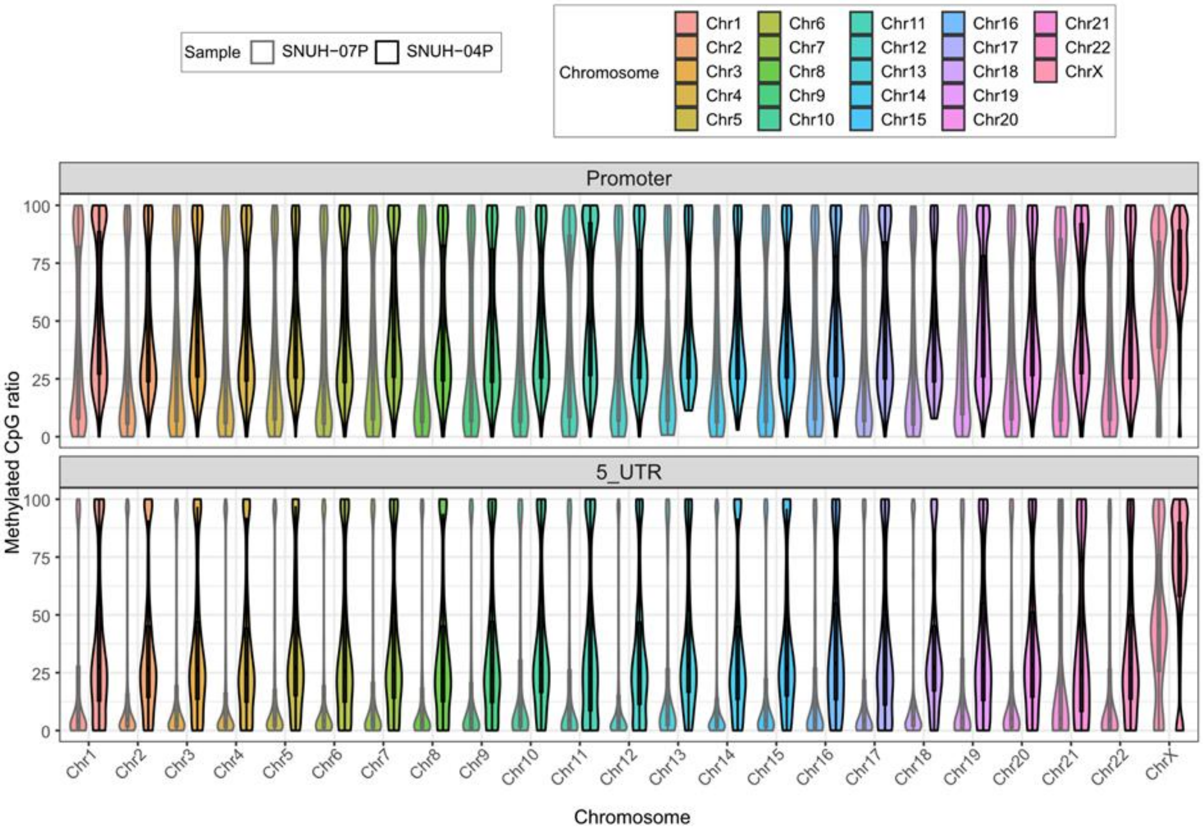


**Figure S3.** Methylation profiles across chromosomes of the patient (SNUH-04P) harboring a *CXXC1* mutation. SNUH-07P is shown as a control sample.

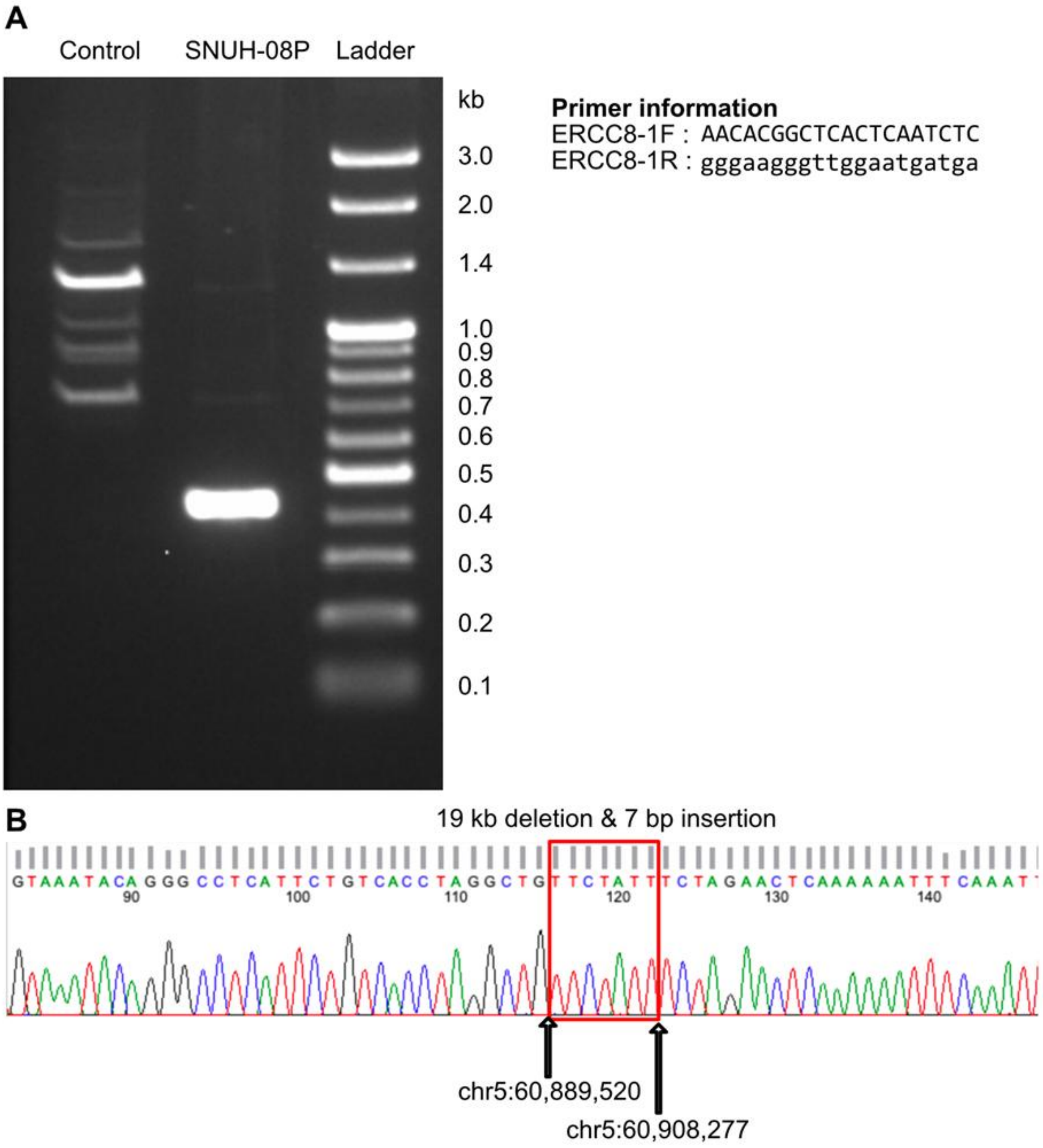


**Figure S4.** Validation of a novel substitution of SNUH-08P identified in *ERCC8*.

**A** PCR validation using a specific primer set. Primer sequences are shown on the right. **B**) Sanger sequencing validation. The red box indicates the 7-bp inserted sequence.

### SUPPLEMENTARY TEXT

#### Diagnosed cases using LRS in addition to previous SRS analyses

#### Case 1. Compound Heterozygous Variant: Unresolved Developmental Disorder

Two siblings (SNUH-09P and SNUH-09S) presented with global developmental delay and characteristic facial dysmorphism suggestive of a familial genetic disorder. Although WES and WGS identified two candidate variants in *SHMT2*, phasing could not be confirmed due to the absence of parental samples. LRS enabled direct phasing and confirmed the variants to be *in trans*.

#### Case 2. Non-Coding Structural Variant: *SLC16A2*-Associated Allan-Herndon-Dudley Syndrome

We recently reported a case study involving two siblings (SNUH-10P and SNUH-10S) with early-onset neurodevelopmental delay, microcephaly, and spasticity. LRS identified a 2.8 kb hemizygous intronic deletion in *SLC16A2*, inherited from their asymptomatic parent. This non-coding deletion encompasses multiple predicted transcription factor binding sites and an eQTL associated with *SLC16A2* expression, and is likely to disrupt regulatory control. This represents the first report of a non-coding deletion associated with Allan-Herndon-Dudley syndrome.

#### Case 3. Complex Insertion: *FKTN*-Associated Congenital Muscular Dystrophy with Precise Breakpoint Resolution by LRS

A patient (SNUH-12P) presented with congenital muscular dystrophy, global developmental delay, hypotonia, intellectual disability, and a myopathic facial appearance. She underwent muscle biopsy showing severe myopathic changes with marked fiber size variation, degeneration/regeneration, and fat infiltration. WGS identified a homozygous complex insertion in chromosome 9 (NC_000009.12:g.105639639_105639640ins[(546);105639640–105639656]); however, the exact structure and breakpoints remained unresolved. LRS enabled precise mapping of the insertion breakpoints and structural characterization of the variant, supporting its role in the disease pathogenesis.
